# Application of a multispecies probiotic reduces gastro-intestinal discomfort and induces microbial changes after colonoscopy

**DOI:** 10.1101/2022.10.18.22281027

**Authors:** J. Labenz, D. P. Borkenstein, F. J. Heil, A. Madisch, U. Tappe, H. Schmidt, B. Terjung, I. Klymiuk, A. Horvath, M. Gross, V. Stadlbauer

## Abstract

Even after decades of research and pharmaceutical development, cancer is still one of the most common causes of death in the western population and the management of cancer will remain a major challenge of medical research. One of the most common types of cancer is colorectal cancer (CRC). Prevention by detection of early-stage precursors is the most reliable method to prevent CRC development. In dependence of age, familial predisposition, and other risk factors the preventative routine screening for CRC by colonoscopy should be performed at least twice in intervals of about ten years. Although colonoscopy is a life-saving clinical examination reducing both incidence and mortality of CRC significantly, it has still a bad reputation in the population as an uncomfortable procedure with unpleasant side effects lasting sometimes over days to weeks. These effects are most likely caused by the bowel preparation before colonoscopy, which is crucial for a successful colonoscopy with high quality. Beside pain, bleeding and other rare but severe complications of colonoscopy, cleaning of the intestinal mucosa alters the gut microbiome significantly and consistently. Abdominal pain, cramps, diarrhea, nausea, bloating, and constipation are common adverse events which can continue to affect patients for days or even weeks after the procedure. In this multicenter, placebo controlled, double blind clinical trial, we investigated the effect of an intervention with a multispecies probiotic formulation for 30 days on the adverse events due to bowel preparation. We show that the treatment of participants with the multispecies probiotic formulation decreases the number of days with constipation significantly, and reduced pain, bloating, diarrhea, and general discomfort. 16S based amplicon analyses reveal recovery of administered probiotic strains from stool samples and differences in alpha diversity dynamics with higher variability in the probiotic group compared to the placebo group. In conclusion, the probiotic ameliorates the side effects after colonoscopy and might be an important supplement to increase acceptance of this life-saving preventative examination. Further, we present here for the first time that probiotic intervention of only 30 days affects alpha diversity parameters in stool samples.

## 1 Introduction

Global cancer statistics exhibit cancer still as the leading cause of death worldwide (1). Tumor entities with the highest prevalence in world population are lung, liver, breast, stomach, and colon cancer (1) with increasing incidences in western populations due to various risk factors like live style, genetic factors, or nutritional habits (2). Colorectal cancer (CRC), if not diagnosed at early stages, is associated with a poor prognosis. Moreover, the incidence of early onset of this tumor form (below the age of 50 years) is increasing during the last 30 years (3). Today, CRC is a global health burden and although therapeutic treatment approaches are improving and five-years survival rates are increasing, a preventive medical examination is still the best way to avoid the development of CRC. The gold standard for CRC-prevention by removal of benign polyps and by early CRC-detection is colonoscopy in asymptomatic individuals (4). Most European countries and a multiplicity of countries worldwide developed preventive programs from age of 50 with ten years of screening intervals (5–7). Colonoscopy is a generally safe and well-tolerated method to visualize the distal terminal ileum and the entire colon (8, 9). Importantly, the quality of this clinical investigation depends largely on an adequate bowel preparation prior to the examination to remove residual fecal content (10, 11). Clinical research aims to improve the methods of bowel preparation with respect to cleansing effect and tolerability (12–14).

Despite all efforts, bowel preparation by colonic lavage is an exhausting procedure for many patients. Standards in bowel preparation define the consumption of oral sulfate solution (OSS) or polyethylene glycol with ascorbic acid (PEG + ASC) (15). During the process of bowel preparation, water passes from the tissue into the lumen of the intestine, thereby flushing fecal content. As side effect of this procedure, the microbial homeostasis might be disturbed and increasing evidence suggests that these preparations have significant impact on the diversity and the composition of the fecal microbiota (16–19). While some studies report short term alterations of the intestinal microbiome (20–22), Drago et al. (18) demonstrated significant and persisting changes regarding the abundance of different bacterial phyla after bowel preparation followed by colonoscopy. The bowel preparation decreased the abundance of *Firmicutes*, especially *Lactobacilli* which have a health-promoting effect, whereas *Proteobacteria* abundance is increased (18,23,24). In addition to the luminal microbiome, it has been shown that standard bowel cleansing adversely affects the mucosa-associated microbiota (25).

Changes in the intestinal microbiome following bowel lavage and colonoscopy may also contribute to abdominal symptoms including bloating, abdominal pain or altered bowel function (26). Even more severe side-effects of the colonoscopy like appendicitis have been reported (27, 28). According to these findings, bowel preparation may induce short-term or even persisting side effects thereby affecting the patient’s wellbeing. A potential therapeutic strategy to restore a balanced composition of the intestinal microbiome after colonoscopy is the use of probiotics – live microorganisms that confer a health benefit on the host when administered in adequate amounts (29). To date, only few studies have evaluated the effect of probiotics on gastrointestinal symptoms and microbiome modulations in patients undergoing colonoscopy and healthy volunteers (30, 31). The administration of probiotic bacteria ameliorates or shortens the days of abdominal pain especially in patients with pain before colon lavage and colonoscopy (30). In a study by Deng et al., the researchers focused on the fecal microbiome showing that bacterial phyla are affected during and after bowel preparation (31). Probiotics administered directly after colon lavage for five to seven days exhibited positive effects on the microbial composition by increasing the abundance of bacterial genera including *Bacteroides* and *Faecalibacterium* and by decreasing the abundance of taxa like *Acinetobacter* and *Streptococcus* (31). These preliminary studies indicated that probiotics may both alleviate side effects arising from bowel preparation followed by colonoscopy and improve intestinal dysbiosis.

In this prospective, double-blind and placebo-controlled study we tested the hypotheses that a multispecies probiotic applied for 30 days immediately after colon lavage followed by a preventive colonoscopy ameliorates duration and intensity of abdominal symptoms and improves the restoration of the intestinal microbiome as assessed by 16S rRNA analysis from of DNA isolated from stool samples.

## 2 Material and Methods

### 2.1 Study overview and design

For this prospective, multicenter, randomized, double-blind and placebo-controlled clinical trial asymptomatic participants undergoing routine colonoscopy for preventative examination were recruited at nine different centers for gastroenterology in Germany. Inclusion criteria for participating were a good health status without diagnosed bowel disease, an age of 50 - 80 years as well as the written consent for participation to the study. Exclusion criteria for participation were diagnosed bowel disease, application of probiotic supplementation in the last four weeks before onset of the study or the use of additional probiotics rather than the study medication. Further, the use of antibiotic therapy in the last four weeks before the colonoscopy or during the 30 days of observation period, immunosuppressive therapy, the use of proton pump inhibitors, an age younger than 50 years or older than 80 years as well as the lack of written, informed consent to participate in the study. We hypothesize in this study that the administration of a special formulated multispecies probiotic rebuilds a reduced intestinal microbiome faster after colonoscopy than administration of a placebo. Strains for the probiotic were selected based on their properties for resistance to acids and pancreatin, adherence to intestinal cells, pathogen inhibition (growth inhibition tests), inhibition of *Clostridioides difficile* growth (toxin production), barrier function (transepithelial electrical resistance test), mast cell inhibition, induction of T-cell differentiation and effect on cytokine production. Further, we hypothesize that the number of days with gastrointestinal complaints are reduced in the group of patients that receives the probiotic formulation compared to the placebo group. To test this hypothesis the primary aim of the study was to examine the influence of a probiotic intervention on the (re-)colonization of the gut microbiome after bowel preparation for colonoscopy. Secondary aims were to determine the number of days with general gastrointestinal complaints (abdominal pain, bloating, heartburn, nausea, vomiting, diarrhea, constipation) the probands reported as well as to the stool characteristics reported from the probands during days after bowel preparation and colonoscopy according to the Bristol Stool scale (BSS) (32). For sample size calculation the ClinCalc samples size calculator was used (https://clincalc.com/Stats/SampleSize.aspx; last accessed September 2022). An anticipated incidence of changes in the intestinal microbial diversity in the probiotic group of 70% and in the placebo group of 35% with a type I error (alpha) of 0.05, and a power of 85% as well as a dropout rate of 15% was estimated. The calculation resulted in a required number of participants of at least 88 probands randomized in a 1:1 ratio to the two intervention groups. The trial was conducted according to the declaration of Helsinki (version 2013). Informed, written consent was obtained from all participants and the study was approved by the responsible ethics committee of the University of Münster (votum number: 2019-201-f-S). All procedures were performed in accordance with the Declaration of Helsinki. The study was registered as clinical trial at the German Register of Clinical Trials (DRKS00018115; https://www.drks.de/drks_web/; accessed 07/2022).

### 2.2 Bowel preparation and colonoscopy

Bowel preparation was performed as split lavage with either Moviprep^®^ or Plenvu^®^ (both Norgine, Amsterdam, The Netherlands) or Eziclen^®^ (Ipsen Pharma GmbH, Germany) according to the manufacturerś instructions. The choice of the product was up to the discretion of the clinician performing the examination. The colonoscopy was performed according to the German quality standards by experienced endoscopists (33). Colonoscopies were performed with CO2 in all cases. Sedation was performed according to the German guidelines for sedation in endoscopy (33). The choice of the sedation drug was up to the discretion of the investigator.

### 2.3 Participants, multispecies probiotic and placebo formulation

Participants were randomized and allocated to each group with the software Research Randomizer^®^ (Urbaniak, G. C., & Plous, S., Version 4.0, retrieved on June 22, 2013, from http://www.randomizer.org/; last access June 2022). The allocation was performed by block randomization at each study center. The probiotic group received the multispecies probiotic formulation containing 3g of *Bifidobacterium bifidum* W23, *Bifidobacterium lactis* W51, *Enterococcus faecium* W54, *Lactobacillus acidophilus* W37, *Lactobacillus rhamnosus* WGG, *Lactococcus lactis* W19 at a concentration of 1×10^9^ cfu/g in a matrix of rice starch, maltodextrin, hydrolyzed rice protein, potassium chloride, manganese sulfate and magnesium sulfate (OMNi BiOTiC^®^ Colonize) twice daily over 30 days. The placebo group received a similar looking, smelling, and tasting powder containing rice starch, maltodextrin, potassium chloride, manganese sulfate and magnesium sulfate. All strains present in the probiotic product have the Qualified Presumption of Safety (QPS) status defined by the European Food Safety Authority (EFSA) (34). Study product packages (probiotic and placebo) were provided with consecutive participant numbers and were blinded to the assignment of the intervention. The randomization code was revealed at the end of the study when all laboratory analyses were completed. During the study, stool samples were collected from the probands at two timepoints. One specimen was taken immediately before colonoscopy related bowel preparation was performed (T1) and the second stool sample 30 days after colonoscopy and treatment with the multispecies probiotic or the placebo (T2).

### 2.4 Sample collection, storage and questionnaires

Stool samples were collected before colonoscopy related bowel preparation and after 30 days of treatment using the Invitek Molecular stool collection tubes with DNA stabilizer (Berlin, Germany) and stored at 4-8°C until nucleic acid extraction and purification according to manufacturer’s recommendations. Total DNA was isolated at the Institute of Clinical Molecular Biology, Christian-Albrechts-University Kiel, Rosalind-Franklin-Str. 12, 24105 Kiel, Germany according to published procedures (35). Briefly, the QIAamp DNA fast stool mini kit automated on the QIAcube (Qiagen, Hilden, Germany) was used for DNA extraction according to manufacturer’s instructions. For mechanic lysis, sample were bead beating using the 0.70 mm Garnet Bead tubes (Dianova, Hamburg, Germany) filled with 1.1 ml InhibitEx lysis buffer in a SpeedMill PLUS (Analytik Jena, Jena, Germany) for 45s at 50Hz according to manufacturer’s instructions and as published (35). Isolated nucleic acids were stored at -20°C until further processing.

### 2.5 Questionnaire

Participants performed a daily questionnaire regarding the occurrence of abdominal pain, bloating, heart burn, nausea, gas, vomiting, diarrhea and constipation for 28 days after colonoscopy. Further, probands provided information on consistency and frequency of their bowel movements based on the BSS (32). Possible frequencies ranked from no stool at all to three times or more often daily. Possible values for stool nature ranked from type 1 (single, solid beads) throughout type 7 (liquid with hardly solid components).

### 2.6 based microbiome analysis

Paired-end sequencing data were received as FASTQ files from the microbiome lab at the Christian-Albrechts-University Kiel according to published procedures (36). Briefly, amplification of the 16S rRNA gene was performed, targeting the hypervariable regions V1-V2 with the target specific primers 27F (5’-AGA GTT TGA TCC TGG CTC AG-3’) and 338R (5’-GCT GCC TCC CGT AGG AGT-3’). Sequencing was executed using the Illumina MiSeq desktop sequencer with 600 cycles version 3 chemistry in 2 x 300bp read mode (Illumina Inc., San Diego, CA, USA). FASTQ raw reads were used for subsequent downstream processing of sequencing data and analysis. Raw reads were uploaded to the European Nucleotide Archive ENA and can be accessed via the project-ID PRJEB56238 (https://www.ebi.ac.uk/ena/submit/webin/; last accessed 09/2022).

### 2.7 Next generation sequencing (NGS) data analysis

FASTQ raw reads were used for data analysis. Subsequent downstream processing of sequencing data was performed using the DADA2 package (https://benjjneb.github.io/dada2/index.html, 1.10) for R (v3.6), adapted to V1-V2 amplicon reads by adjusting the filterAndTrim () paramters: truncLen=c(230, 180), trimLeft=c(5, 5), truncQ=5, rm.phix=T. Number of bases for error inference were set to 10⁹. Resulting amplicons sequence variants (ASVs) were taxonomically annotated using the Genome Taxonomy Database (GTDB; https://gtdb.ecogenomic.org/) release 202 reference files using the Bayesian classifier of the DADA2 package (assignTaxonomy()). Sequence data were rarefied to 9,000 per sample and ASVs collapsed into genus-level taxonomic bins based on the taxonomic annotation. ASVs not annotated at genus level were collapsed into respective higher order taxonomic bins.

Alpha diversity measures Shannon diversity (vegan::diversity()), Chao1 (vegan::estimateR()) and richness were calculated on the rarefied genus level count abundance tables. Group-wise comparisons in alpha diversity were performed using Wilcoxon rank sum test and Wilcoxon signed rank test for independent and repeated measurements, respectively.

Bray-Curtis dissimilarity was calculated to assess beta diversity. Differences in community composition and dispersion were assessed by permutational multivariate analysis of variance (vegan::adonis()) with 10,000 permutations performed to assess significance.

Analysis of taxon-specific abundance differences between treatment groups and timepoints were based on CLR-transformed count data to account for the compositional nature of microbiome data with pseudocount=1 added to each respective count. CLR-transformed values < 0 were set to 0. For between group comparisons, linear models were used to assess differences in the zero-truncated CLR-transformed abundances. For comparisons between sampling time points, linear mixed models (nlme::lme()) were used with individuals as random effects variables. Individuals were included in the analysis of a taxonomic group if the respective clade was present in the samples of that individual at least in one timepoint.

Multivariable associations were examined using MaAsLin2, a method based on Compound Poisson Linear Models (CPLM) specialized for compositional, zero-inflated data sets. Within this method the unmodified feature table was subjected to trimmed mean of M-values (TMM) normalization. A combination of treatment and timepoint was used as fixed effect (37).

Additionally, analysis on beta diversity and probiotic strain recovery were performed after initial quality check, primer removal and truncation of reads at 250 bases (forward) and 200 (reverse) reads, respectively. Maximum expected error was set to 2 for both, forward and reverse reads. Error rates were modelled based on 451457 reads from 14 samples for the forward reads and 576611 reads from 18 samples for reverse reads. Data were de-replicated and introduced to the dada2 inference algorithm. ASVs were merged and filtered for chimera and ultra-low abundant sequence variants. A naïve Bayesian classifier, Ribosomal Database Project (RDP) classifier, implemented in the dada2 assignTaxonomy function was used to classify the sequence variants into higher-order taxonomy based on the SILVA V132 database (38). Species annotation was allowed at 100% identity. The web-based application OmicsNet 2.0 (https://www.omicsnet.ca/; accessed 07/2022) was used to extract possible metabolite networks around the features of interest (39). The potential score which indicates the probability that a given taxon produces a metabolite was set to 90%, current, universal and metabolites without pathway annotation, were excluded from analysis.

### 2.8 Gastrointestinal symptoms data analysis

All statistics were done using R version 4.1.3 (2022-03-10) (40) as well as GraphPad Prism version for Windows (GraphPad Software, La Jolla California USA, www.graphpad.com). The number of days with intestinal symptoms were compared between groups using Mann-Whitney-u-tests and unpaired samples t-test, the proportion of patients reporting symptoms within the groups were compared using chi-square tests. Distance-based redundancy analysis (dbRDA) was performed to assess the association between the gastrointestinal symptoms and the change in the microbiome composition in both groups. An OTU table was prepared and centered log ratio-transformed using pldist::data_prep() with the argument paired=TRUE and pldist::pltransform(), respectively. Gower’s distance was calculated using pldist() and distance based redundancy analysis was performed with vegan::dbrda(). GraphPad Prism version 9.3.1 for Windows (GraphPad Software, La Jolla California USA, www.graphpad.com) was used for data visualization according to manufacturer’s instructions.

## 3 Results

All participants included into the study were enrolled between September 2019 and July 2021. In total 91 participants were included to the study and randomized to probiotic and placebo intervention groups with a dropout of four patients who did not keep the appointment for colonoscopy or withdrew their informed consent (Figure 1). Out of the remaining 87 patients, age (average age=59.5 years), sex (52% male, 48% female) and body mass index (BMI) (average BMI=26.61 kg/m^2^) (Table 1) matched probands who underwent colonoscopy, 45 participants were allocated to the probiotic group and 42 to the placebo group as shown in Figure 1. From the remaining 87 patients in total 76 (n=39 for probiotic and n=37 for placebo) provided stool samples at both time points (T1 just before colonoscopy and T2 after 30 days of treatment intervention with the placebo and the probiotic). 36 probands in the probiotic group and 35 probands in the placebo group completed the questionnaire on gastrointestinal discomfort and were included to the statistical analysis. In addition to information on abdominal pain, bloating, heartburn, nausea, vomiting, diarrhea, and constipation the subjects provided information about the nature and frequency of their bowel movements according to BSS.

**Figure 1:**
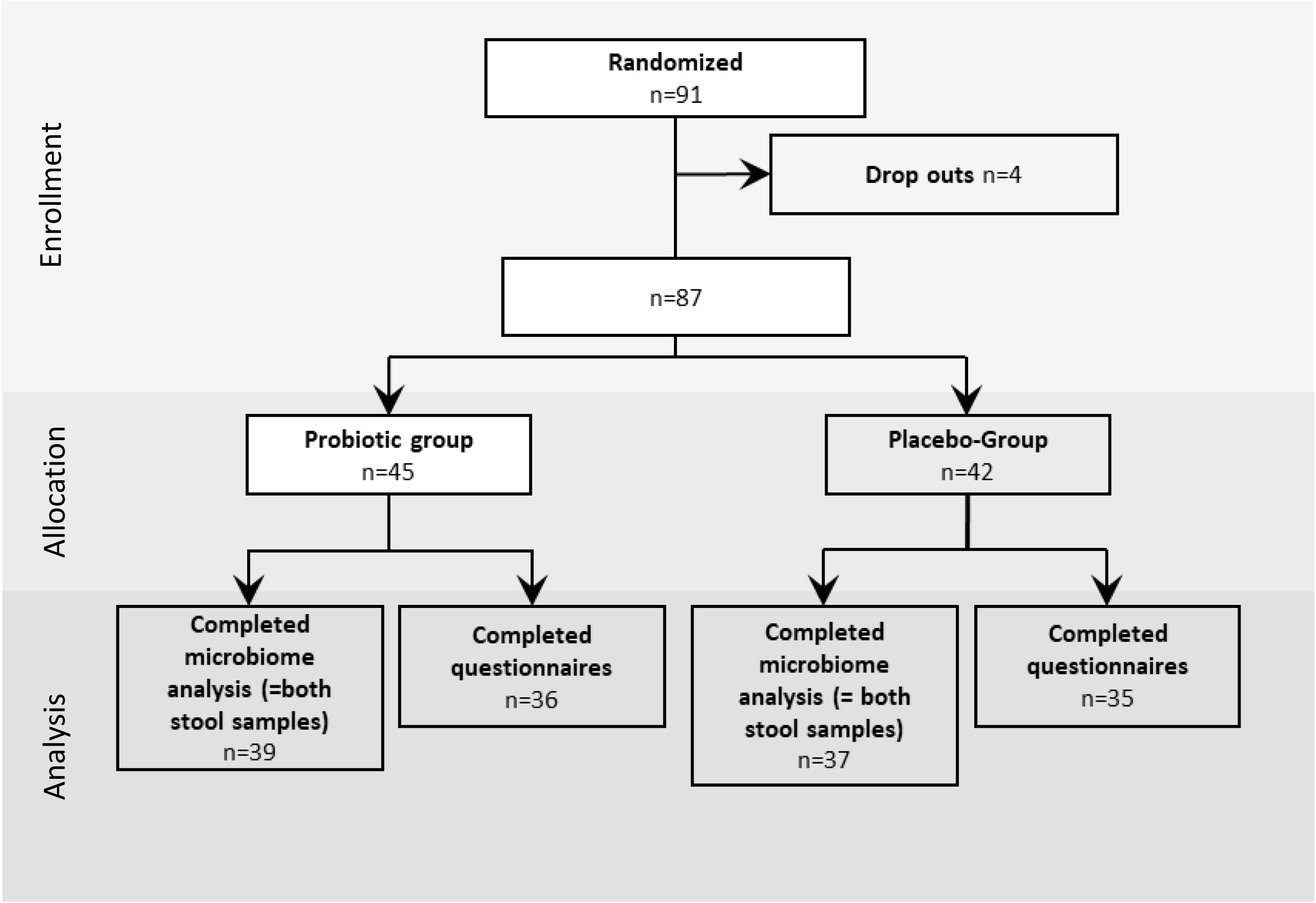
Study design: Enrollment, allocation and analysis of proband questionnaires data and stool samples of the study. The intervention was performed for 30 days after colonoscopy, the questionnaires was performed for 28 days.

**Table 1:**
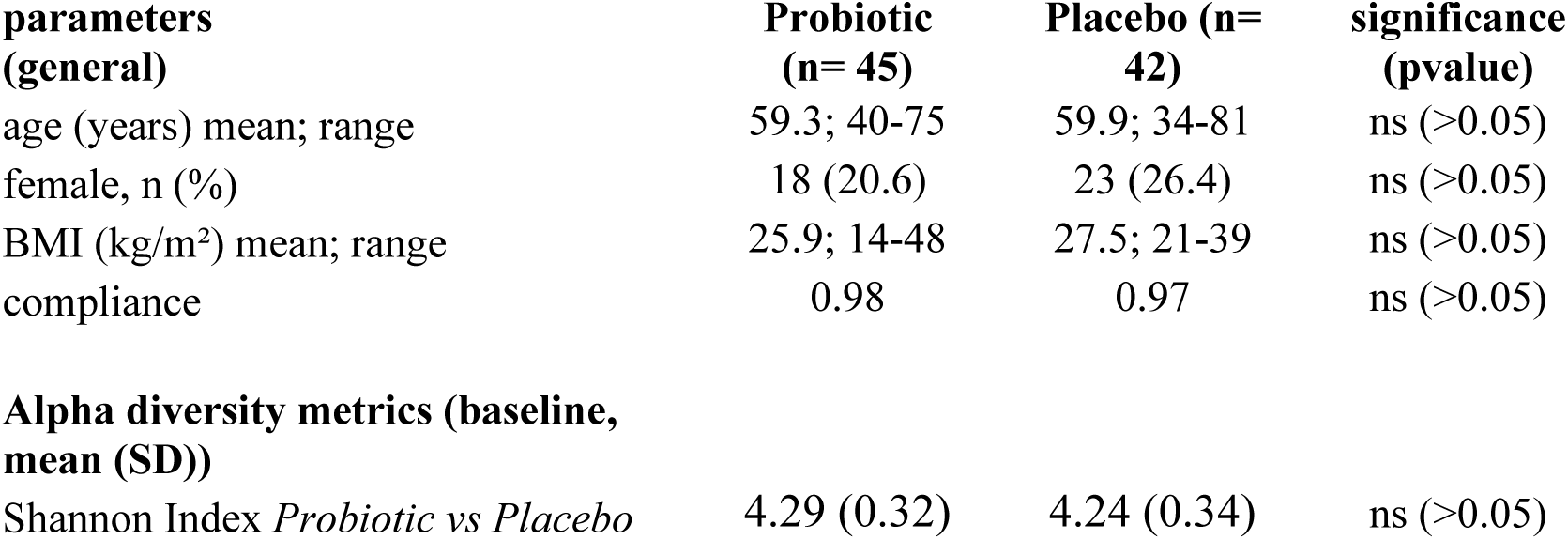
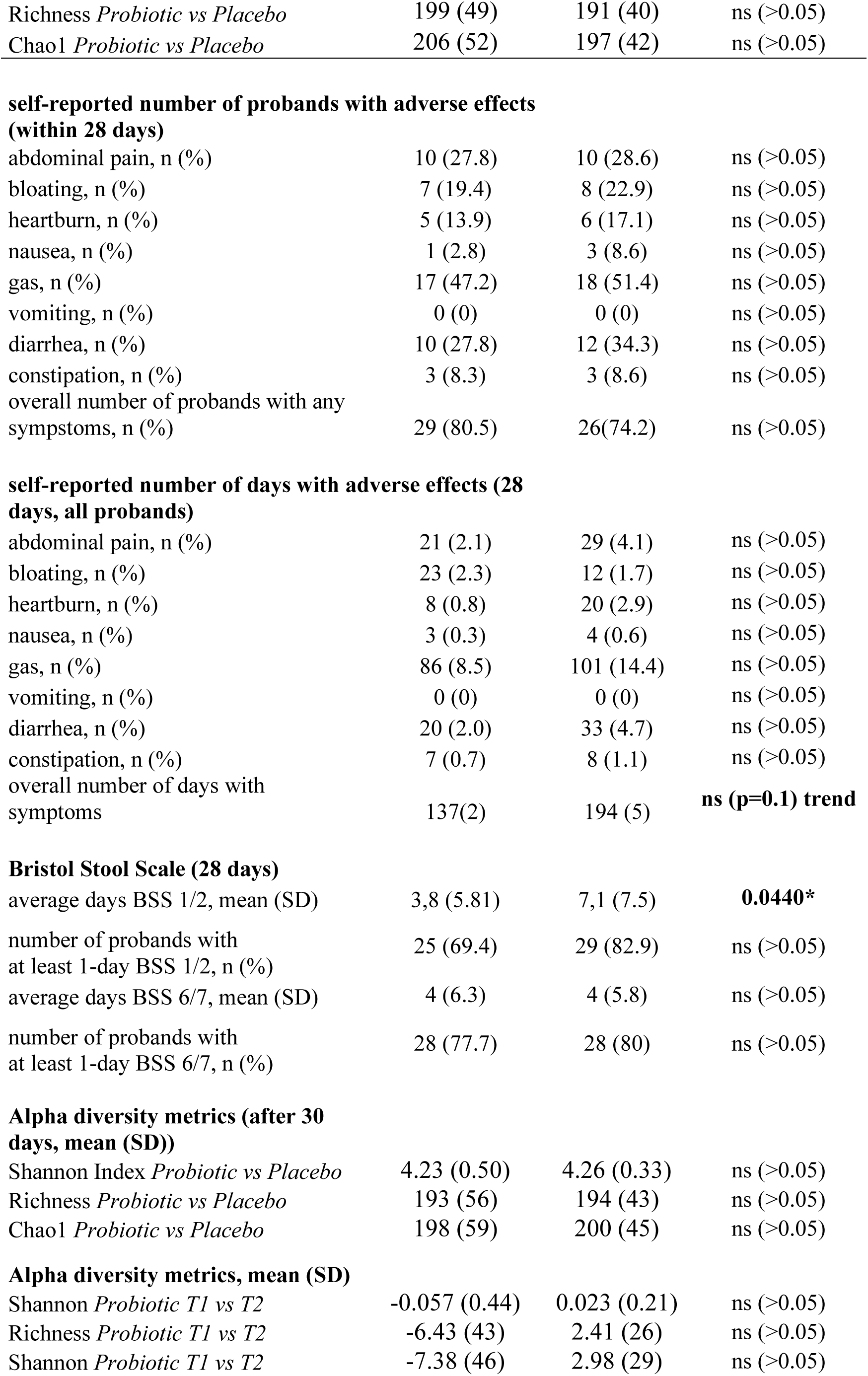
Patient characteristics: Baseline characteristics and after 30 days of treatment and 28 days of questionnaire. At baseline the probiotic and placebo groups do not differ in any parameter analyzed. ns…not significant.

| parameters<br>(general) | Probiotic<br>(n= 45) | Placebo (n= 42) | significance<br>(pvalue) |
| --- | --- | --- | --- |
| age (years) mean; range | 59.3; 40-75 | 59.9; 34-81 | ns (>0.05) |
| female, n (%) | 18 (20.6) | 23 (26.4) | ns (>0.05) |
| BMI (kg/m<sup>2</sup>) mean; range | 25.9; 14-48 | 27.5; 21-39 | ns (>0.05) |
| compliance | 0.98 | 0.97 | ns (>0.05) |
| **Alpha diversity metrics (baseline, mean (SD))** |  |  |  |
| Shannon Index *Probiotic vs Placebo* | 4.29 (0.32) | 4.24 (0.34) | ns (>0.05) |
| Richness *Probiotic vs Placebo* | 199 (49) | 191 (40) | ns (>0.05) |
| Chao1 *Probiotic vs Placebo* | 206 (52) | 197 (42) | ns (>0.05) |
| **self-reported number of probands with adverse effects (within 28 days)** |  |  |  |
| abdominal pain, n (%) | 10 (27.8) | 10 (28.6) | ns (>0.05) |
| bloating, n (%) | 7 (19.4) | 8 (22.9) | ns (>0.05) |
| heartburn, n (%) | 5 (13.9) | 6 (17.1) | ns (>0.05) |
| nausea, n (%) | 1 (2.8) | 3 (8.6) | ns (>0.05) |
| gas, n (%) | 17 (47.2) | 18 (51.4) | ns (>0.05) |
| vomiting, n (%) | 0 (0) | 0 (0) | ns (>0.05) |
| diarrhea, n (%) | 10 (27.8) | 12 (34.3) | ns (>0.05) |
| constipation, n (%) | 3 (8.3) | 3 (8.6) | ns (>0.05) |
| overall number of probands with any<br>symptoms, n (%) | 29 (80.5) | 26(74.2) | ns (>0.05) |
| **self-reported number of days with adverse effects (28 days, all probands)** |  |  |  |
| abdominal pain, n (%) | 21 (2.1) | 29 (4.1) | ns (>0.05) |
| bloating, n (%) | 23 (2.3) | 12 (1.7) | ns (>0.05) |
| heartburn, n (%) | 8 (0.8) | 20 (2.9) | ns (>0.05) |
| nausea, n (%) | 3 (0.3) | 4 (0.6) | ns (>0.05) |
| gas, n (%) | 86 (8.5) | 101 (14.4) | ns (>0.05) |
| vomiting, n (%) | 0 (0) | 0 (0) | ns (>0.05) |
| diarrhea, n (%) | 20 (2.0) | 33 (4.7) | ns (>0.05) |
| constipation, n (%) | 7 (0.7) | 8 (1.1) | ns (>0.05) |
| overall number of days with<br>symptoms | 137(2) | 194 (5) | **ns (p=0.1) trend** |
| **Bristol Stool Scale (28 days)** |  |  |  |
| average days BSS 1/2, mean (SD) | 3,8 (5.81) | 7,1 (7.5) | **0.0440\*** |
| number of probands with<br>at least 1-day BSS 1/2, n (%) | 25 (69.4) | 29 (82.9) | ns (>0.05) |
| average days BSS 6/7, mean (SD) | 4 (6.3) | 4 (5.8) | ns (>0.05) |
| number of probands with<br>at least 1-day BSS 6/7, n (%) | 28 (77.7) | 28 (80) | ns (>0.05) |
| **Alpha diversity metrics (after 30 days, mean (SD))** |  |  |  |
| Shannon Index *Probiotic vs Placebo* | 4.23 (0.50) | 4.26 (0.33) | ns (>0.05) |
| Richness *Probiotic vs Placebo* | 193 (56) | 194 (43) | ns (>0.05) |
| Chao1 *Probiotic vs Placebo* | 198 (59) | 200 (45) | ns (>0.05) |
| **Alpha diversity metrics, mean (SD)** |  |  |  |
| Shannon *Probiotic T1 vs T2* | -0.057 (0.44) | 0.023 (0.21) | ns (>0.05) |
| Richness *Probiotic T1 vs T2* | -6.43 (43) | 2.41 (26) | ns (>0.05) |
| Shannon *Probiotic T1 vs T2* | -7.38 (46) | 2.98 (29) | ns (>0.05) |

### 3.1 Microbiome analysis – primary aim

16S rRNA gene-based microbiome analysis was performed from 39 participants that received the multispecies probiotic and from 37 participants that received placebo after colonoscopy, respectively. Before the start of bowel cleansing, there was no statistically significant differences in Shannon alpha diversity index calculation at genus level of stool samples between the two randomized groups (probiotic and placebo) (Figure 2A). Spearman’s rank correlation analysis revealed no statistically significant relation between Shannon diversity index at genus level and the body mass index (BMI) (Figure 2B). Nevertheless, there was a significant correlation (p-value=0.042, rho=-0.232) of reduced Shannon diversity index at genus level with increasing age (Figure 4C). Before start of the treatment in all baseline samples analyzed (T1) the five most abundant genera were *Faecalibacterium*, *Bacteroides*, *Prevotella*, *Phocaeicola* and *Alistipes* (Supplementary Table 1). All normalized read counts of bacterial taxa at T1 and T2 from phylum to genus level analysis for all samples are shown in supplementary table 1.

**Figure 2:**
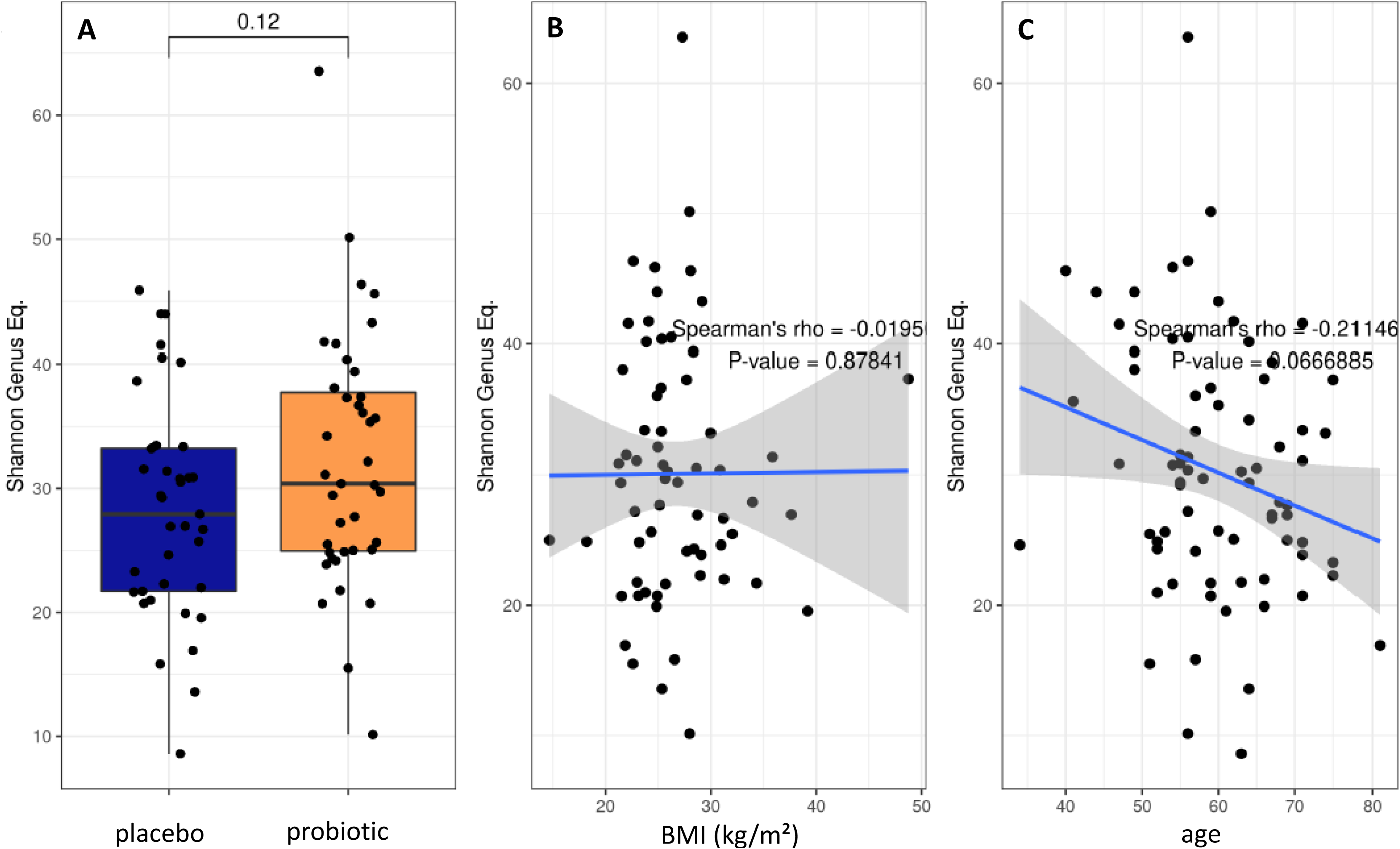
Baseline characteristics of the two proband groups. (A) Box blot analysis of Shannon diversity index for alpha diversity calculation at genus level at baseline before start of bowel preparation and treatment. Spearman’s rank correlation test on the effect of (B) BMI and (C) Age on alpha diversity.

After baseline characterization of the samples that ensured the equality of the study groups, alpha and beta diversity calculations were performed for both proband groups (probiotic and placebo) between T1 and T2. Alpha diversity metrics including Shannon diversity index, Chao1 and richness and did not reveal statistically significant differences before and after 30 days of treatment with either the placebo or the probiotic (p>0.05) (Figure 3A-C). The effective number of species detected in the samples was calculated with Shannon number equivalent (Figure 3D). Although, the Shannon number equivalent did not change significantly neither in the probiotic nor in the placebo group between the two timepoints T1 and T2 tested (Figure 3D), the delta of this value changed significantly between the probiotic and the placebo group after four weeks of treatment (p=0.036) (Figure 3E) suggesting that absolute changes in alpha diversity between timepoints are possibly larger in the probiotic group.

**Figure 3:**
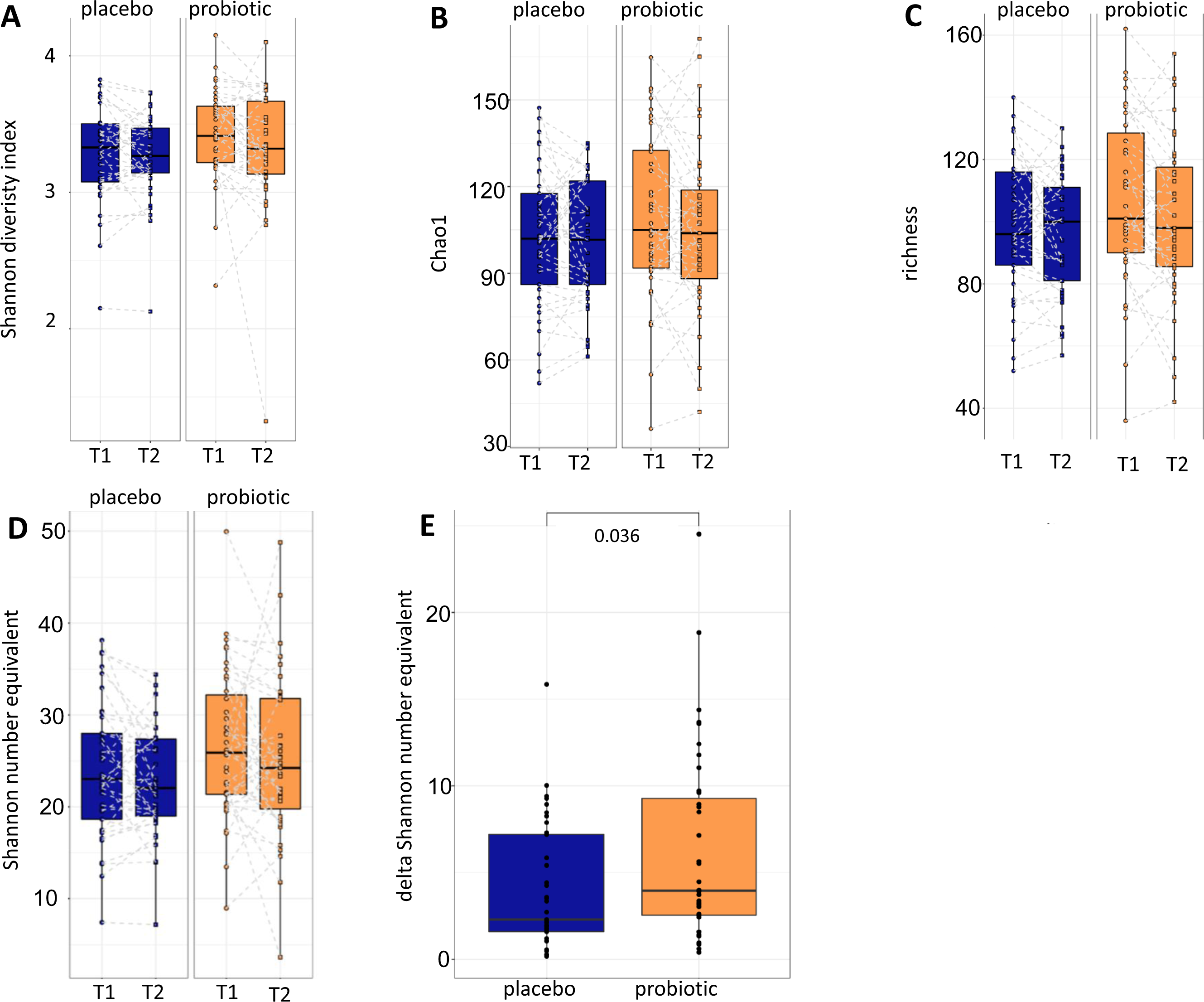
Alpha diversity calculations. (A) Shannon diversity index, (B) Chao1 and (C) richness between T1 and T2 of the placebo and the probiotic group. Additionally, the (D) Shannon number equivalent and the (E) delta of the Shannon number equivalent are calculated. T1…timepoint 1, T2…timepoint 2.

Metric multidimensional scaling (MDS) analysis using Bray-Curtis dissimilarity as beta diversity metric was performed for the placebo and the probiotic group before (T1) and after (T2) 30 days of treatment. No statistically significant differences were found between the comparisons analyzed (p>0.05) (Supplementary Figure 1A). Further, the comparison between T1 and T2 for the placebo and the probiotic group separately, did not result in statistically significant regulations, either (Supplementary Figure 1B and C). Additionally, the community level changes calculated as the delta of Bray-Curtis dissimilarity values between the two time points did not alter significantly between the two treatment groups (p=0.27; data not shown).

#### 3.1.1 Differential taxa abundance

Finally, we performed mixed-effects model of centered and log-ratio transformed (CLR) abundances analysis of all taxa that occurred in at least 10 samples with a relative abundance of at least 0.5% across all samples between timepoints and groups. Abundance of the genus *Clostridioides sp*. CAG:417 decreased in the placebo group between T1 and T2 (p<0.05) but increased in the probiotic group from T1 to T2 (p<0.05) (Figure 4A). The taxon Bacillales bacterium UBA660 decreased in the probiotic group between T1 and T2 significantly (p<0.05), without significant changes in the placebo group (p>0.05) (Figure 4B). Further taxa that exhibited significant changes were *Mesosutterella* and CAG.354 that increased in T2 in the probiotic as well as in the placebo group compared to T1 (p<0.05, data not shown) and an uncultured Firmicutes UMGS1491 (*Oscillospiraceae*) (p<0.05, data not shown) that was more abundant in the probiotic group at both time points compared to the placebo group. The genera *Duodenibacillus*, *Ruminiclostridium* and an uncultured *Clostridioides sp*. (Firmicutes) UMGS1663 were decreased in their relative abundance in the probiotic group independent of treatment and time point (p>0.05, data not shown). No taxonomic groups were significantly associated with any of the variables (group, timepoint) after correction for multiple testing.

**Figure 4:**
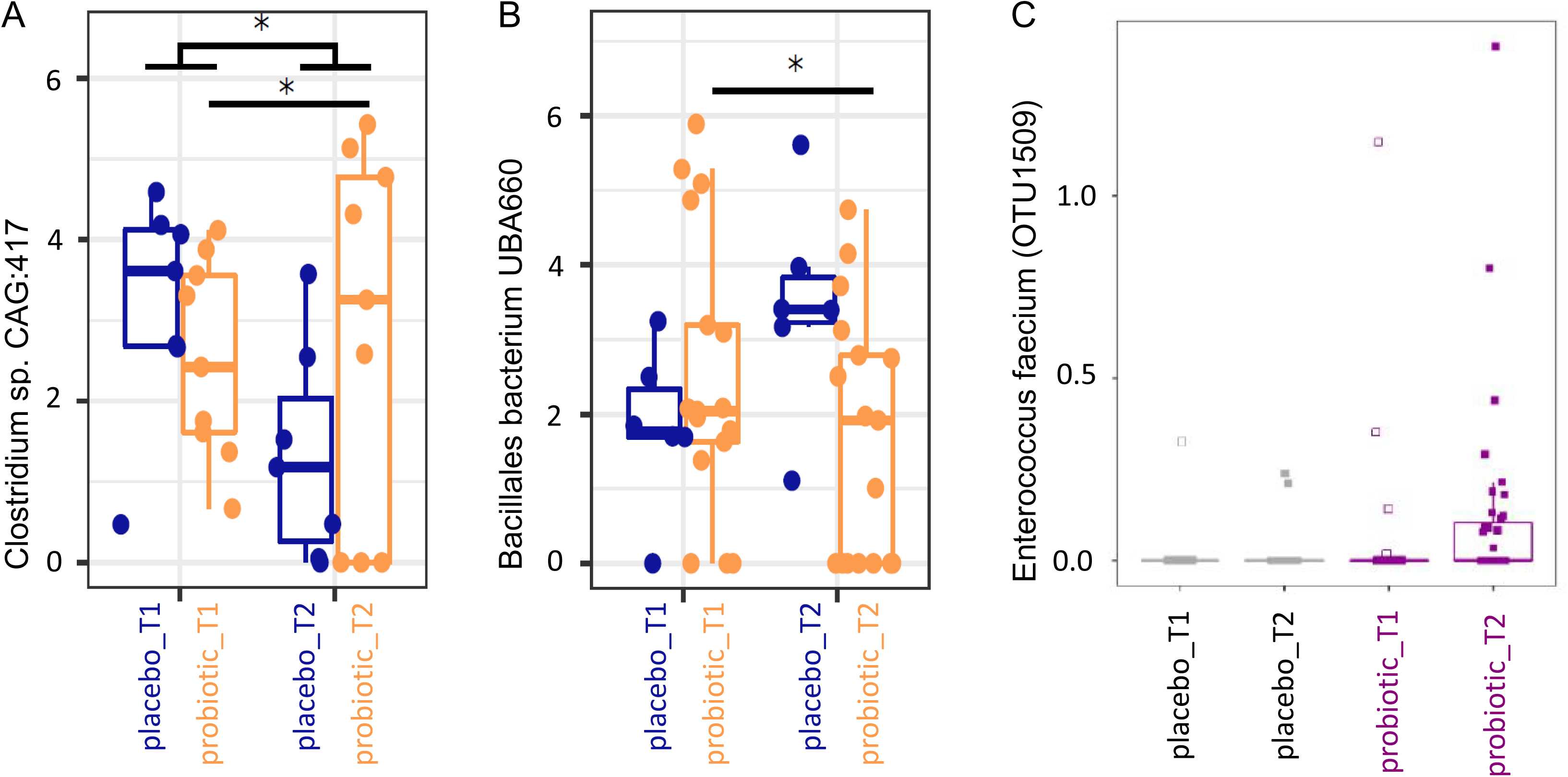
(A-B) Box plot analysis of mixed model analysis of centred and log-ratio transformed (CLR) abundances. Taxa with relative abundances of at least >0.5% in at least 10 samples across all timepoints were included. *p<0.05 in the mixed model analysis. (C) Differential abundance in percent of Enterococcus faecium determined by MaAsLin2 between the two time points for the probiotic and the placebo group.

At T2 no statistically significant correlations were found with Spearman’s rank correlation between the time points and the parameters age and body mass index (p>0.05).

#### 3.1.2 Probiotic recovery and prediction analysis on metabolomic pathways

The bacterial strains in the multispecies probiotic formulation were Bifidobacterium bifidum W23, Bifidobacterium lactis W51, Enterococcus faecium W54, Lactobacillus acidophilus W37, Lactobacillus rhamnosus WGG, Lactococcus lactis W19 in equimolar ratios. The 16S rRNA based analyses revealed recovery of reads derived from nucleic acids corresponding to the applied probiotic strains according to table 2. In the probiotic group all applied strains were recovered from the stool analysis in at least one sample with at least one copy (table 2). Further, MaAsLin2 analysis revealed Enterococcus faecium to be significantly associated with probiotic treatment at T2 (p<0.05, relative abundance) (Figure 4C). One Bacteroides species increased in the probiotic group (p<0.05, relative abundance). Based on these two species (Enterococcus faecium and Bacteroides ovatus) we performed predicted metabolomics analysis and drew a knowledge-based metabolite network. The resulting network had 17 nodes (i.e. metabolites) and 27 edges (i.e. connections between the nodes) (Supplementary Figure 2). Based on the likely metabolites of the input bacteria, the functions phenylalanine-, tyrosine- and tryptophan biosynthesis were significantly associated with these bacteria, with four metabolites shared between them (2-Dehydro-3-deoxy-D-arabino-heptonate 7-phosphate, 3-Dehydroshikimate, Shikimate and 5-O-(1-Carboxyvinyl)-3-phosphoshikimate). All four metabolites were located in the early stages of the pathway, also called ‘Shikimate-pathway’ (Supplementary Figure 2).

**Table 2:**
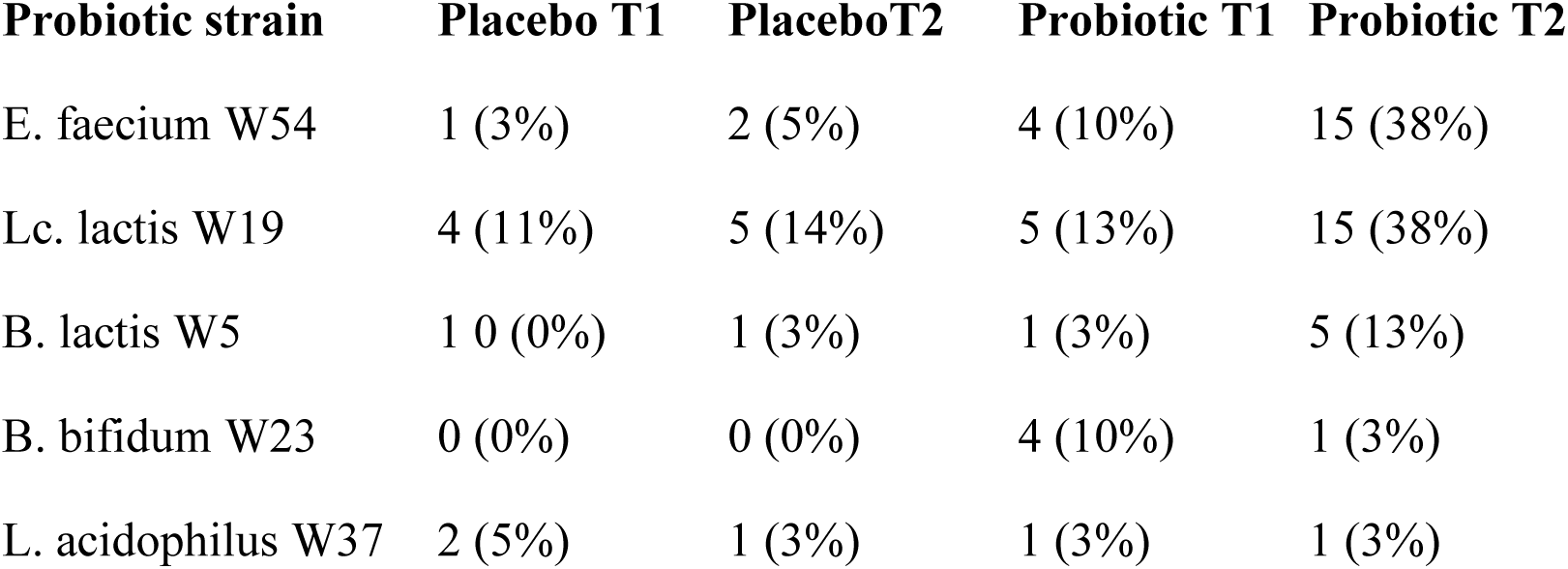
Probiotic recovery in all groups and timepoints. Data are given as number (percentage) of samples with at least one copy of the respective sequence. Number of samples (percentage of samples).

### 3.2 Gastrointestinal symptoms – secondary aim

The average data from questionnaires of the probands and their adverse events reported after colonoscopy are provided in Table 1. Common adverse events of the participants of this study after colonoscopy were abdominal pain, bloating, heartburn, nausea, gas, vomiting, diarrhea and constipation.

We observed a significant reduction of the number of days with constipation in the probiotic group compared to the placebo group (p=0.044) (Figure 5A). Days with general intestinal complaints revealed a non-significant trend (p>0.05) towards a decreased number (Figure 5B) and the number of days with bloating were also reduced (Figure 5C) (p>0.05). In total, out of the 72 probands that completed the questionnaires, 69 patients experienced days with symptoms during an observational period of 30 days. The change in overall days of symptoms however did not reach statistical significance between groups (p=0.1) (Figure 3).

**Figure 5:**
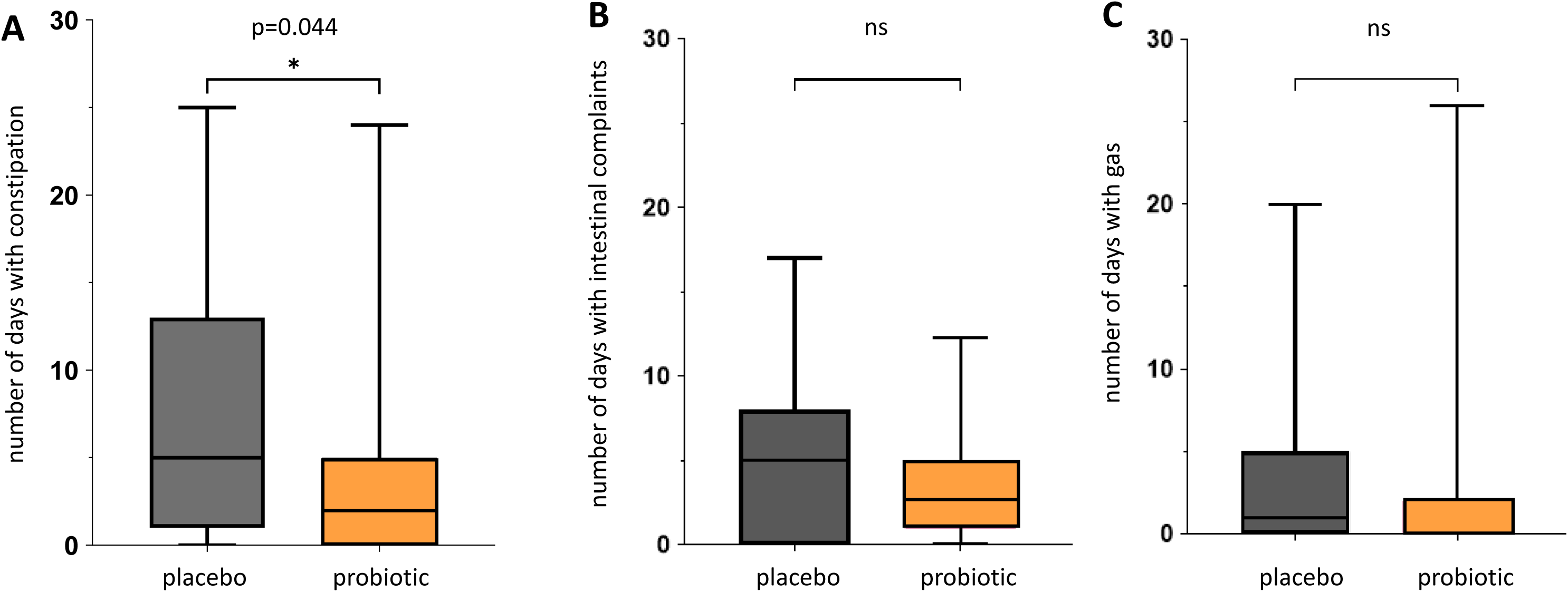
Questionnaire for gastrointestinal symptoms after colonoscopy. Days spent with gastrointestinal symptoms for placebo and probiotic group. (A) Number of days self-reported with constipation according to Bristol stool scale. (B) Number of days self-reported with intestinal complaints. (C) Number of days self-reported with bloating. The intestinal complaints include all recorded adverse events for all days of observation. ns…not statistically significant.

**Figure 6:**
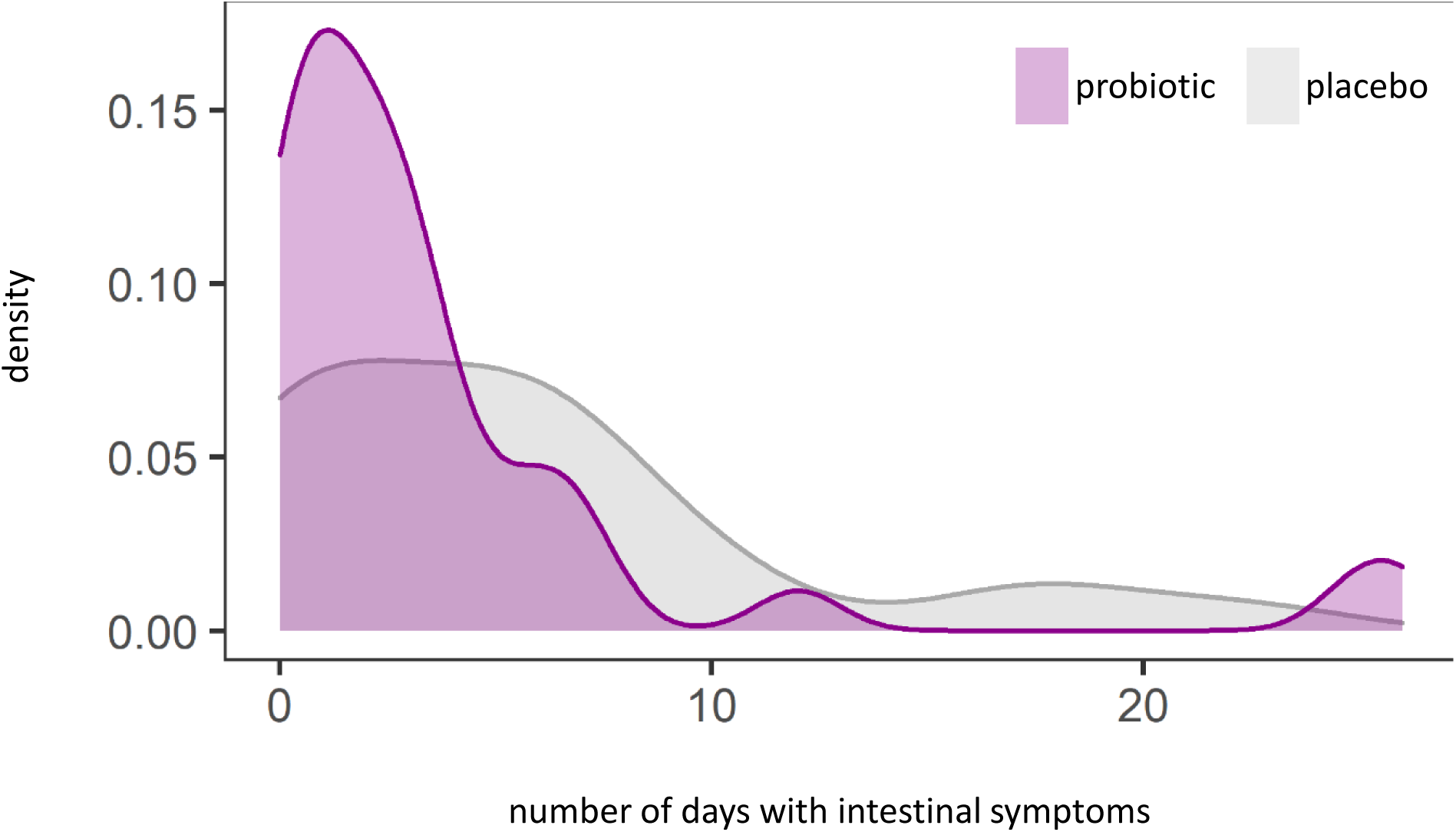
Density plot analysis of general intestinal symptoms (abdominal pain, bloating, heartburn, nausea, vomiting, diarrhea, constipation) (p=0.1).

Distance-based redundancy analysis (dbRDA) revealed no statistically significant association of the self-reported gastrointestinal symptoms with changes in the 16S based bacterial microbiome during the intervention period for any further gastrointestinal complaints analyzed (abdominal pain, heartburn, nausea, vomiting, diarrhea) (data not shown).

## 4 Discussion

During this clinical trial the primary aim was to compare the intestinal microbiome immediately before a colonoscopy and the microbiome after 30 days of administration of the multispecies probiotic OMNi-BiOTiC® Colonize. We hypothesized that the intestinal microbiome detected by 16S analyses from stool samples will change by application of the multistrain probiotic compared to a placebo. Our results clearly show a higher delta in the Shannon number equivalent in the probiotic group compared to the placebo indicating more alterations in the expected number of species calculated by this value between T1 and T2 in the probiotic compared to the placebo group. Therefore, the primary aim of the study defined as the characterization of the changes in stool microbial pattern was reached.

The secondary aims of this clinical trial were the analysis of abdominal complaints between the experimental groups and the classification of the stool. We hypothesized less abdominal complaints and less constipation or diarrhea in the probiotic compared to the placebo group. The secondary aim was reached with a significant reduction in the number of days with constipation according to the BSS and a non-significant trend of less abdominal complaints in the probiotic group compared to the placebo group.

In a prospective, double blind, placebo controlled and randomized interventional study we could therefore show for the first time that a multispecies probiotic formulation has a positive impact on side effects occurring after screening colonoscopy and its related bowel preparation and on procedure induced disturbances of the intestinal microbiome in healthy probands.

Post-colonoscopy induced symptoms are often abdominal pain, bloating, diarrhea, or constipation ((7,26,41). A recent publication reports bloating in 25% of the analyzed patients as well as abdominal pain and discomfort in 5%-11% of the analyzed cases, as the most common adverse effects after colonoscopy (41). A prospective cohort study with a total of 502 patients reported minor complications in 34% of probands. Most common adverse effects were bloating with 25% and abdominal pain with 11% (42) corroborating the findings from other studies (43). In our study cohort, 80% of the patients without abdominal symptoms before the procedure claimed abdominal symptoms at any of the 30 days after the colonoscopy. Flushing of the intestine is on the one hand a strong mechanical load for the gut and the intestinal mucus. On the other hand, the beneficial microbiota becomes imbalanced by this massive intervention in the intestinal community (21, 24). Although, minor and interim side effects of colonoscopy are often of little medical interest, we argue that the good tolerability of colonoscopy is an important prerequisite to convince as many people as possible from the important and lifesaving screening colonoscopy (26, 44).

In the study presented here, we could show a significant reduction in days with constipation and a trend towards reduced days with bloating and diarrhea after colonoscopy in the probiotic treated group. This is in concordance with previous studies on the effect of probiotics on colonoscopy related side effects (30,31,45). Earlier studies analyzed the effect of single strain formulations only. This is the first time a multispecies probiotic formulation with six selected strains was tested to prevent gastro-intestinal complaints after colonoscopy. Based on the data derived from our study, a sample size calculation is possible to confirm our clinical concept of reduction of post-colonoscopy GI complaints by a multispecies probiotic.

We know from previous studies that the intestinal microbiome is reduced massively upon colonoscopy preparation and these alterations in the bacterial community might be an important factor for the occurrence of adverse events after the treatment (21, 24). Although other alpha diversity calculations did not reveal statistical significance in our study, the delta of Shannon number equivalent changed significantly between the two sample groups. The Shannon number equivalent gives the number of equally likely states needed to produce the given Shannon diversity index, so this is the effective number of elements and minimizes the risk of missing changes in the diversity. Our results indicate an increased variation of expected microbial alpha diversity in the probiotic group comparing the sample before colonoscopy and after 30 days of intervention. The biological meaning of this finding needs to be further explored in the future. A weakness of the study is that no intermediate points were taken for microbiome analyses to characterize the time course of microbial changes compared between the interventional groups.

Analyzing the recovery rates of the used bacterial strains we found all the species from the probiotic formulation in the stool samples of participants in the probiotic group, however, a large proportion of patients showed no recovery of probiotic strains at all. Considering that 50% of the human stool is suspected to be microbiota (46), we hypothesize that the bacterial load of the stool samples was too high, and the coverage used during 16S based next generation sequencing analysis too low to recover the probiotic strains from all the patients’ samples, that received the probiotic formulation. We can also not fully exclude non-compliance with the study product although this was part of the questionnaire (Table 1). Especially, the significantly higher abundance of *Enterococcus faecium* in stool samples of the second time point in the probiotic group suggests that the supplemented probiotic strains not only survive the gastric passage but also shows proliferation. Although we are not able to differentiate between live and dead microorganism with the used method, we hypothesize that the supplemented cell number would be far too low for detection in stool biomass. The low recovery rate of the probiotic strains of the genus *Bifidobacterium sp*. might result from the used primers for target amplification, as 27F and 338R primers are known to lack the detection of *Bifidobacterium sp*. (47). Further clinical studies with increased numbers of participants and primers for target specific 16S rRNA amplification might probably improve the significance of the results.

A weakness of the study is that we observed a dropout rate of about 20% of probands who did not complete the daily stool questionnaire. As we calculated with a dropout rate of 15% we have not reached the desired number of evaluable questionnaires (n=88). Subjects did not explain their dropout, but we hypothesize that the burden of the detailed questionnaires was too much for them. Therefore, higher dropout rates especially for long-term microbiome studies with assessment of patient reported outcomes need to be considered.

In conclusion we demonstrated in a randomized, placebo controlled, double-blind study conducted in a homogenous population undergoing screening colonoscopy that the application of a multispecies probiotic at a concentration of 2×10^9^ cfu/g with 6 g per day starting right after colonoscopy, ameliorates side effects of colonoscopy induced constipation significantly. Moreover, we observed a trend towards a lower load of other gastrointestinal symptoms and we found changes in the gut microbiome composition that may be associated with health (48). These clinically relevant findings should be confirmed in a large-scale randomized trial.

## 5 Figures

## 6 Tables

## 8 Author Contributions

JL, DPB, FJH, AM, UT, HS, BT, IK, AH, MG, VS designed and performed the experiments, analyzed the data, and wrote the manuscript.

JL, DPB, FJH, AM, UT, HS, BT, IK, AH, MG, VS analyzed data, contributed materials and reviewed the manuscript.

JL, DPB, FJH, AM, UT, HS, BT, IK, AH, MG, VS provided expertise and feedback and wrote and reviewed the manuscript.

## Data Availability

Raw reads were uploaded to the European Nucleotide Archive ENA and can be accessed via the project-ID PRJEB56238 (https://www.ebi.ac.uk/ena/submit/webin/; last accessed 09/2022).

## Acknowledgments

We thank the probands for their participation to this study.

## 12 Supplementary Material

**Supplementary Table 1:** Rarefied read numbers of microbial taxa from phylum to genus level of stool samples before colonoscopy and after 30 days of application of a multispecies probiotic or a placebo.

**Supplementary Figure 1:** Bray-Curtis dissimilarity plot for Beta diversity analysis of both groups and all time points (A) and for the placebo group and the probiotic group between T1 and T2, respectively. Data points are colored by intervention group and shapes indicate timepoints (circle: T1, square: T2). MDS…metric multidimensional scaling.

**Supplementary Figure 2:** Knowledge-based metabolite networks based on Enterococcus faecium and Bacteroides ovatus.

## 13 Data Availability Statement

**Supplementary Figure 1:**
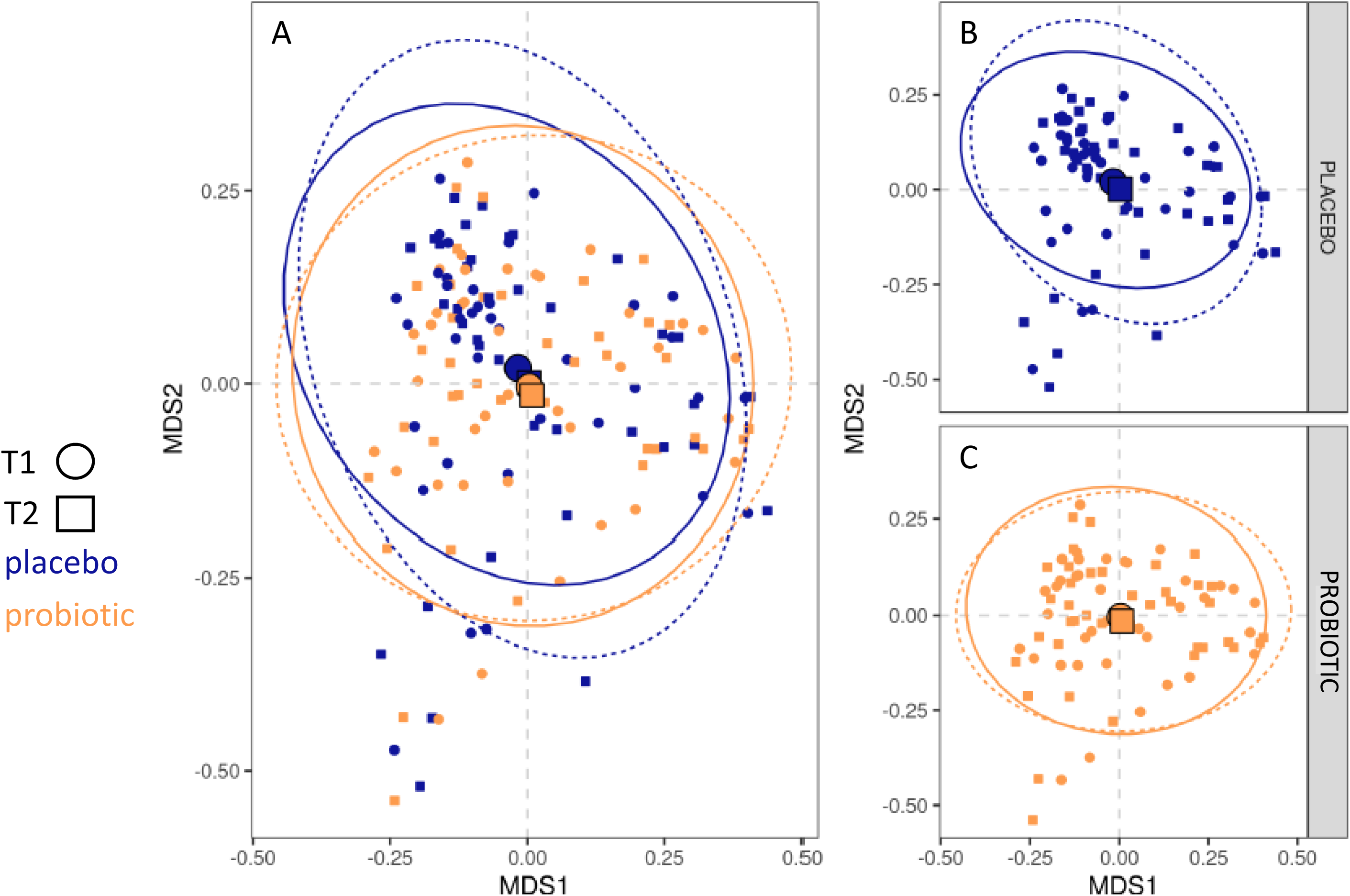
Bray-Curtis dissimilarity plot for Beta diversity analysis of both groups and all time points (A) and for the placebo group and the probiotic group between T1 and T2, respectively. Data points are colored by intervention group and shapes indicate timepoints (circle: T1, square: T2). MDS…metric multidimensional scaling.

**Supplementary Figure 2:**
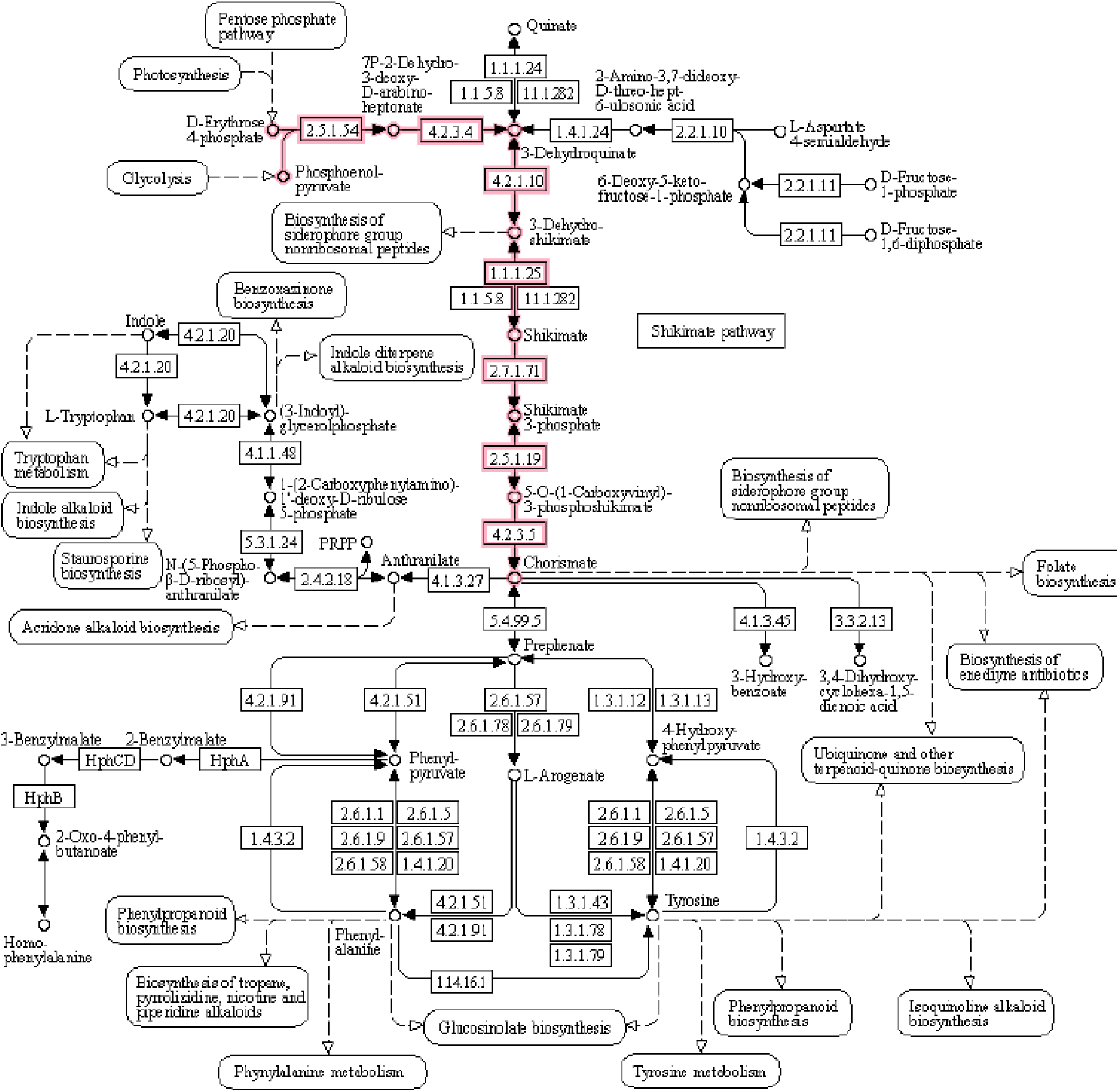
Knowledge-based metabolite networks based on *Enterococcus faecium* and *Bacteroides ovatus*.

